# REACT-1 round 6 updated report: high prevalence of SARS-CoV-2 swab positivity with reduced rate of growth in England at the start of November 2020

**DOI:** 10.1101/2020.11.18.20233932

**Authors:** Steven Riley, Kylie E. C. Ainslie, Oliver Eales, Caroline E. Walters, Haowei Wang, Christina Atchison, Claudio Fronterre, Peter J. Diggle, Deborah Ashby, Christl A. Donnelly, Graham Cooke, Wendy Barclay, Helen Ward, Ara Darzi, Paul Elliott

## Abstract

**Background:** England is now in the midst of its second wave of the COVID-19 pandemic. Multiple regions of the country are at high infection prevalence and all areas experienced rapid recent growth of the epidemic during October 2020.

**Methods:** REACT-1 is a series of community surveys of SARS-CoV-2 RT-PCR swab-positivity in England designed to monitor the spread of the epidemic and thus increase situational awareness. Round 6 of REACT-1 commenced swab-collection on 16th October. A prior interim report included data from 16th to 25th October for 85,971 participants. Here, we report data for the entire round on 160,175 participants with swab results obtained up to 2nd November 2020.

**Results:** Overall weighted prevalence of infection in the community in England was 1.3% or 130 people per 10,000 infected, up from 60 people per 10,000 in the round 5 report (18th September to 5th October 2020), doubling every 24 days on average since the prior round. The corresponding R number was estimated to be 1.2. Prevalence of infection was highest in North West (2.4%, up from 1.2%), followed by Yorkshire and The Humber (2.3% up from 0.84%), West Midlands (1.6% up from 0.60%), North East (1.5% up from 1.1%), East Midlands (1.3% up from 0.56%), London (0.97%, up from 0.54%), South West (0.80% up from 0.33%), South East (0.69% up from 0.29%), and East of England (0.69% up from 0.30%). Rapid growth in the South observed in the first half of round 6 was no longer apparent in the second half of round 6. We also observed a decline in prevalence in Yorkshire and The Humber during this period. Comparing the first and second halves of round 6, there was a suggestion of decline in weighted prevalence in participants aged 5 to 12 years and in those aged 25 to 44 years. While prevalence remained high, in the second half of round 6 there was suggestion of a slight fall then rise that was seen nationally and also separately in both the North and the South.

**Conclusion:** The impact of the second national lockdown in England is not yet known. We provide here a detailed description of swab-positivity patterns at national, regional and local scales for the period immediately preceding lockdown, against which future trends in prevalence can be evaluated.

## Introduction

England is now in the second wave of the COVID-19 pandemic [1]. Since May 2020, we have been carrying out near real-time surveillance of the epidemic in England through successive rounds of the REal-time Assessment of Community Transmission-1 (REACT-1) study, based on RT-PCR of self-administered swabs [2–7]. We recently reported a rapidly rising prevalence of infections during the first half of round 6, with a doubling time of 9.0 (6.1, 18) days, for swabs collected between 16th and 25th October 2020 [7]. Here, we report results for the complete period of data collection in round 6. This includes swabs obtained up until 2nd November 2020, three days before England entered a second national lockdown [8].

## Methods

We have described REACT-1 methods elsewhere [3]. Briefly, we are using RT-PCR to analyse self-administered nose and throat swabs (or parent/guardian administered for children ages 5 to 12 years) obtained from random samples of the population of England from the age of five years upwards. Swabs were maintained on a cold chain until analysed in a single laboratory. The sample was designed to obtain approximately equal numbers of people in each of the 315 lower-tier local authorities (LTLAs) in England, using the National Health Service (NHS) list of GP registered patients to obtain the sample. We have aimed for a sample size of between 120,000 and 160,000 people for each round of data collection, with response rates varying between 22% and 31%.

Once participants provided their swab they were invited to complete a brief health and lifestyle questionnaire. SARS-CoV-2 prevalence (with 95% confidence intervals) was estimated nationally and regionally and by various socio-demographic characteristics, e.g. age. Both unweighted and weighted estimates were obtained, the latter by adjusting for region, deprivation and ethnicity, as well as differential response, so as to be representative of the population in England. We analysed time-trends in swab positivity both between and within rounds using exponential growth and decay models and we carried out multivariable logistic regression to investigate associations of socio-demographic variables and symptoms with swab positivity.

As well as quantifying regional trends, we have investigated SARS-CoV-2 prevalence at sub-regional level, using a geospatial model. The goal of these analyses was to estimate the England-wide geographical variation in the swab-positive prevalence *P(x)* where *x* is a lower layer super output area (LSOA) population-weighted centroid. We model the log-odds of *P(x)* as the sum of a region-level mean and an unobserved, zero-mean spatially correlated stochastic process *S(x)* that captures local variation around each region-wide mean.

Conditional on *P(x)* the LSOA-level numbers of positive swab tests are independent and binomially distributed. We estimate the model parameters by Monte Carlo maximum likelihood, then draw samples from the joint predictive distribution of *P(x)* over all LSOAs, i.e. the joint distribution conditional on the observed numbers of positive swab tests. We then scale-up to LTLA-level by converting the LSOA-level samples to LTLA-level population-weighted averages. All computations used the PrevMap package [9] within the R computing environment [10].

We obtained research ethics approval from the South Central-Berkshire B Research Ethics Committee (IRAS ID: 283787).

## Results

We found 1,732 positives from 160,175 swabs giving an unweighted prevalence of 1.08% (95% CI, 1.03%, 1.13%) and a weighted prevalence of 1.30% (1.21%, 1.39%) (Table 1). The weighted prevalence estimate was more than double that of 0.60% (0.55%, 0.71%) obtained in the prior round 5 of the study [4].

**Table 1.** Unweighted and weighted prevalence of swab-positivity across six rounds of REACT-1.

| Round | Tested swabs | Positive swabs | Unweighted prevalence (95% CI) | Weighted prevalence (95% CI) | First sample | Last sample |
| --- | --- | --- | --- | --- | --- | --- |
| 1 | 120,620 | 159 | 0.13% (0.11%, 0.15%) | 0.16% (0.13%, 0.19%) | 1/5/2020 | 1/6/2020 |
| 2 | 159,199 | 123 | 0.077% (0.065%, 0.092%) | 0.088% (0.068%, 0.11%) | 19/6/2020 | 7/7/2020 |
| 3 | 162,821 | 54 | 0.033% (0.025%, 0.043%) | 0.040% (0.027%, 0.053%) | 24/7/2020 | 11/8/2020 |
| 4 | 154,325 | 137 | 0.089% (0.075%, 0.11%) | 0.13% (0.096%, 0.15%) | 20/8/2020 | 8/9/2020 |
| 5 | 174,949 | 824 | 0.47% (0.44%, 0.50%) | 0.60% (0.55%, 0.71%) | 18/9/2020 | 5/10/2020 |
| 6** | 160,175 | 1732 | 1.08% (1.03%, 1.13%) | 1.30% (1.21%, 1.39%) | 16/10/2020 | 2/11/2020 |
| 6a** | 85,971 | 863 | 1.00% (0.94%, 1.07%) | 1.28% (1.16%, 1.42%) | 16/10/2020 | 25/10/2020 |
| 6b** | 74,210 | 869 | 1.17% (1.10%, 1.25%) | 1.32% (1.20%, 1.45%) | 26/10/2020* | 2/11/2020 |

\*Includes small number of samples from prior period
\*\*Total tested swabs for round 6 is less than the sum of rounds 6a and 6b because of participant withdrawal.

The increase in prevalence between this study and the prior round 5 represents a national doubling time of 24 (22, 27) days with a corresponding R estimate of 1.19 (1.17,1.21) (Figure 1, Table 2). This R estimate from sequential rounds is similar to that reported for the prior round 5 alone of 1.16 (1.05, 1.27). We do not report an R estimate for the current round 6 overall because, using maximum likelihood logistic regression, we found strong evidence in favour of a non-linear prevalence function during this period (ΔAIC > 50, Table 3).

**Table 2.** Estimates of growth rate, doubling time and reproduction number for rounds 5 and 6 together and for rounds 6a and 6b alone.

| Data used | Outcome | Growth rate $r$ (1/days) | R number | $p(R>1)$ | Doubling (+) / halving (-) time (days) |
| --- | --- | --- | --- | --- | --- |
| Rounds 5 and 6 | All positives | 0.029 ( 0.026 , 0.032 ) | 1.19 ( 1.17 , 1.21 ) | 1.000 | 24.0 ( 26.5 , 21.8 ) |
|  | Non-symptomatics | 0.030 ( 0.026 , 0.035 ) | 1.20 ( 1.17 , 1.23 ) | 1.000 | 22.9 ( 26.5 , 20.0 ) |
|  | Positive for both E and N genes | 0.025 ( 0.021 , 0.028 ) | 1.16 ( 1.14 , 1.19 ) | 1.000 | 28.0 ( 32.4 , 24.7 ) |
|  | Positive for both E and N genes or positive only for N gene with CT 35 or less | 0.028 ( 0.025 , 0.031 ) | 1.19 ( 1.16 , 1.21 ) | 1.000 | 25.0 ( 28.0 , 22.5 ) |
| Round 6a | All positives | 0.077 ( 0.040 , 0.115 ) | 1.56 ( 1.27 , 1.89 ) | 1.000 | 9.0 ( 17.3 , 6.0 ) |
|  | Non-symptomatics | 0.138 ( 0.082 , 0.191 ) | 2.10 ( 1.60 , 2.65 ) | 1.000 | 5.0 ( 8.4 , 3.6 ) |
|  | Positive for both E and N genes | 0.054 ( 0.006 , 0.099 ) | 1.38 ( 1.04 , 1.75 ) | 0.987 | 12.8 ( 114.8 , 7.0 ) |
|  | Positive for both E and N genes or positive only for N gene with CT 35 or less | 0.057 ( 0.017 , 0.097 ) | 1.40 ( 1.11 , 1.73 ) | 0.997 | 12.1 ( 41.7 , 7.1 ) |
| Round 6b | All positives | -0.024 ( -0.047 , -0.001 ) | 0.85 ( 0.73 , 0.99 ) | 0.020 | -28.4 ( -14.8 , * ) |
|  | Non-symptomatics | -0.074 ( -0.110 , -0.039 ) | 0.59 ( 0.43 , 0.77 ) | <0.001 | -9.3 ( -6.3 , -17.9 ) |
|  | Positive for both E and N genes | 0.000 ( -0.028 , 0.028 ) | 1.00 ( 0.83 , 1.18 ) | 0.507 | * ( -24.9 , 25.2 ) |
|  | Positive for both E and N genes or positive only for N gene with CT 35 or less | -0.009 ( -0.034 , 0.016 ) | 0.95 ( 0.80 , 1.11 ) | 0.250 | -79.9 ( -20.2 , 42.8 ) |

\* Doubling and halving times not given when growth rates are very close to 0.

**Table 3.** Comparison of parsimony of maximum likelihood linear and smooth maximum likelihood logistic regression models of daily swab positivity.

| Round | AIC(linear) -<br>AIC(smooth)* |
| --- | --- |
| 1 | 2.3 |
| 2 | 1.1 |
| 3 | 0.0 |
| 4 | 7.3 |
| 5 | 0.0 |
| 6 | 54.8 |

\* Akaike information criterion

**Figure 1.**
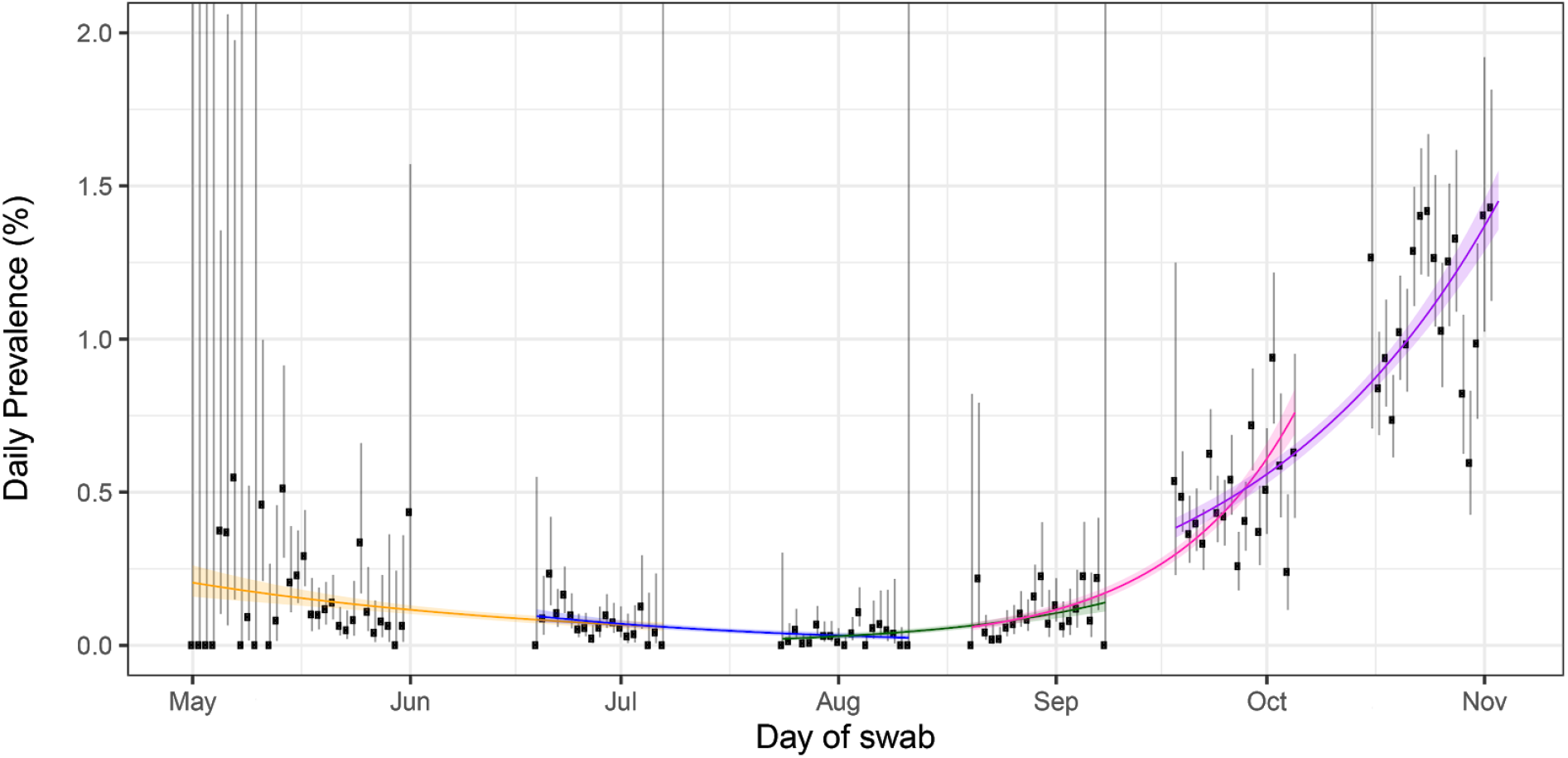
Constant growth rate models fit to REACT-1 data for sequential and individual rounds. Points show unweighted prevalence estimate and vertical lines show 95% binomial confidence intervals (CIs). Because there were few swabs taken on some days, some upper bounds to CIs are truncated. Models fit to REACT-1 data for sequential rounds; 1 and 2 (yellow), 2 and 3 (blue), 3 and 4 (green), 4 and 5 (pink), and 5 and 6 (purple).Shaded areas show 95% credible intervals. Note that of the 932,171 swab tests only 918,543 had a date and so were included in the temporal analysis (3,008 of 3,029 positives).

Following the rapid growth reported during the first half of round 6 [7], here we describe little evidence of growth during the second half. Weighted prevalence for the earlier part of round 6 [7] was 1.28% (1.16%, 1.42%) compared with 1.32% (1.20%, 1.45%) for the most recent data with estimates of R of 1.56 (1.27, 1.89) and 0.85 (0.73, 0.99) for each half of round 6 respectively. We investigated the possibility of systematic differences in the participant characteristics between the two halves of round 6 in comparison with the first and second halves of rounds 4 and 5. While we did find differences in characteristics within rounds, patterns were similar across rounds 4, 5 and 6 (not shown).

There appears to be additional temporal structure in the most recent round 6 data not captured by the average trend described above. A p-spline [11] model suggests a fall and then rise in prevalence during this most recent period, with the lowest point around October 30th (Figure 2). A similar pattern was seen when fitting separate curves to data for the North (including Midlands) and the South (Figure 3). Similar patterns were also seen when holding out each region in turn and refitting the p-spline (Figure 4).

**Figure 2.**
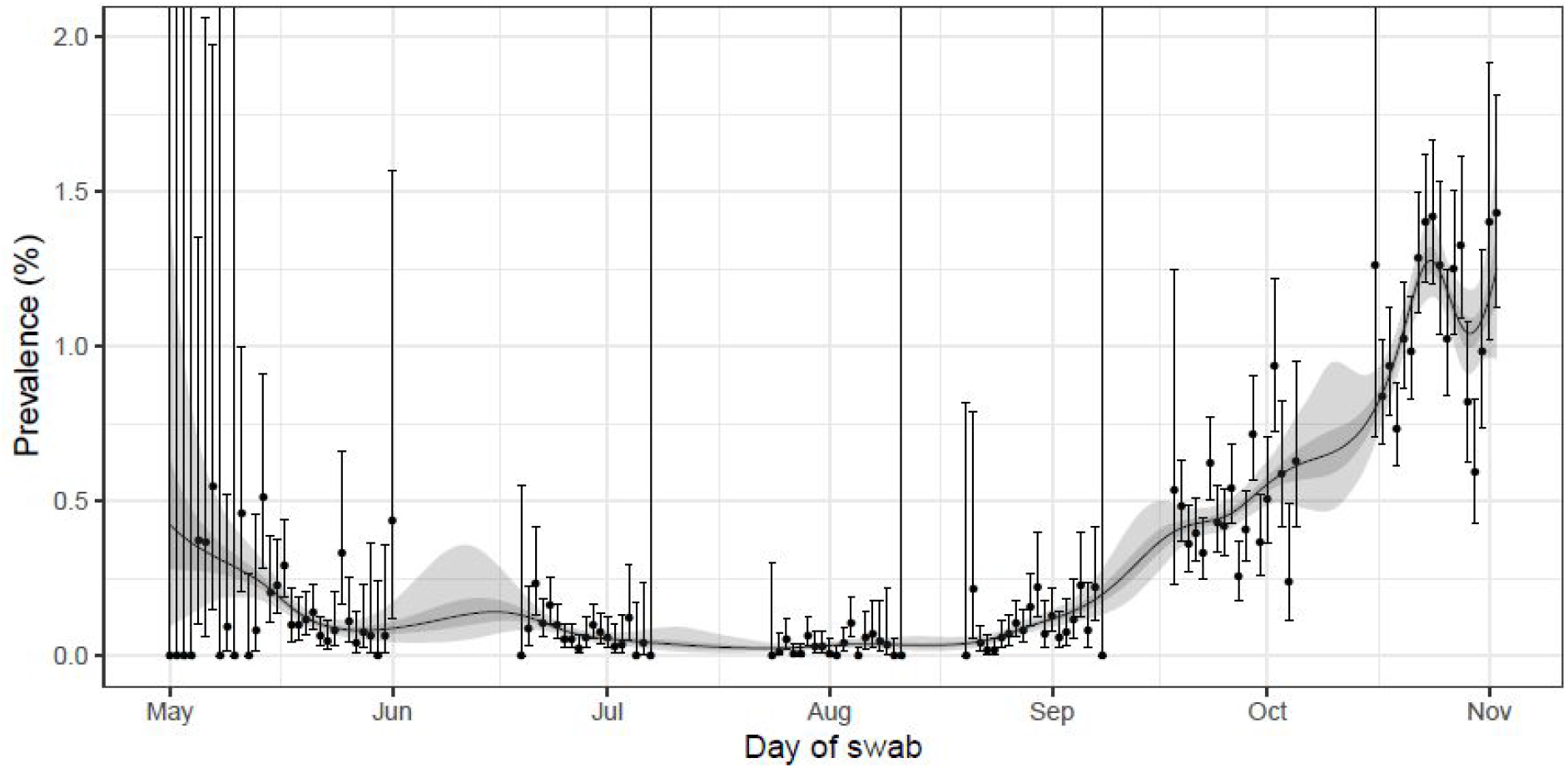
Prevalence of swab-positivity estimated using a p-spline for the full period of the study with central 50% and 95% posterior credible intervals. Points show unweighted prevalence estimate and vertical lines show 95% binomial confidence intervals (CIs). Because there were few swabs taken on some days, some upper bounds to CIs are truncated. Note that of the 932,171 swab tests only 918,543 had a date and so were included in the temporal analysis (3,008 of 3,029 positives).

**Figure 3.**
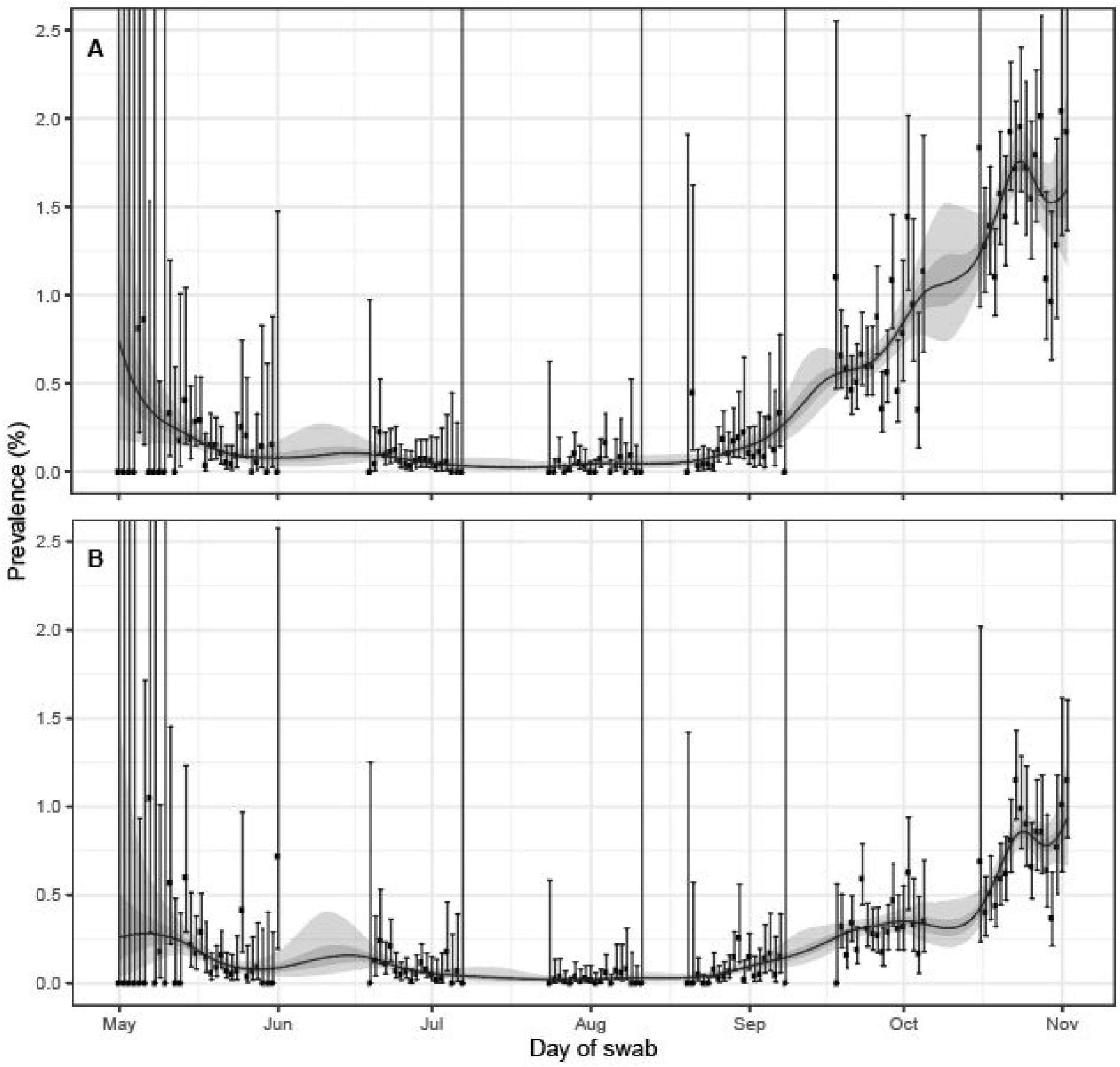
Prevalence of swab-positivity estimated using a p-spline for the full period of the study with central 50% and 95% posterior credible intervals. Points show unweighted prevalence estimate and vertical lines show 95% binomial confidence intervals (CIs). Because there were few swabs taken on some days, some upper bounds to CIs are truncated. **A** The model fit to a subset of the data containing only the regions North East, North West, Yorkshire and The Humber, East Midlands and West Midlands. **B** The model fit to a subset of the data only containing the regions London, East of England, South East and South West. Note that of the 932,171 swab tests 918,543 had a date and so were included in the temporal analysis (3,008 of 3,029 positives).

**Figure 4.**
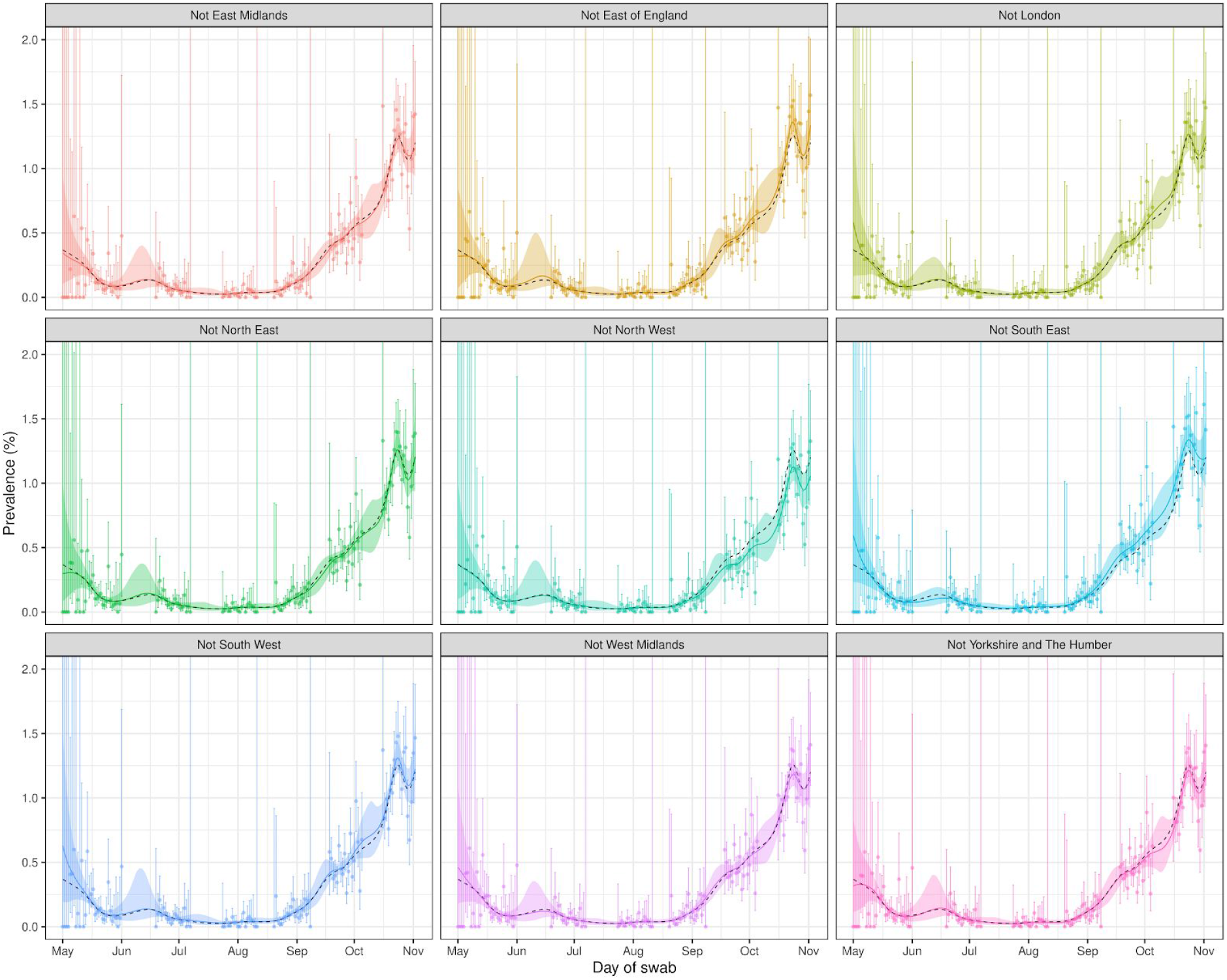
Prevalence of swab-positivity estimated using a p-spline for the full period of the study with central 95% posterior credible intervals for subsets of the data leaving out one region at a time from the analysis. The black dashed line shows the p-spline fit to all available data. Points show unweighted prevalence estimate and vertical lines show 95% binomial confidence intervals (CIs). Because there were few swabs taken on some days, some upper bounds to CIs are truncated. Note that of the 932,171 swab tests 918,543 had a date and so were included in the temporal analysis (3,008 of 3,029 positives).

At regional level, based on analysis of rounds 5 and 6 together, R was increasing (Figure 5, Table 4, Table 5a). However, consistent with the national trend, the rapid growth in the South during the first half of round 6 was no longer apparent in the more recent data (Table 4, Figure 6). While the highest prevalence in the first half of round 6 was in Yorkshire and The Humber, since then, there has been a decline in prevalence in that region (Figure 6). However, the fall in Yorkshire and The Humber in the most recent period does not explain the national pattern (Figure 4).

**Table 4.** Estimates of growth rate, doubling time and reproduction number for rounds 5 and 6 together and for rounds 6a and 6b for english regions.

| Data used | Region | Growth rate $r$ (1/days) | R number | $p(R>1)$ | Doubling (+) / halving (-) time (days) |
| --- | --- | --- | --- | --- | --- |
| Rounds 5 and 6 | South East | 0.035 ( 0.027 , 0.043 ) | 1.24 ( 1.18 , 1.29 ) | 1.000 | 19.8 ( 25.4 , 16.1 ) |
|  | North East | 0.019 ( 0.008 , 0.030 ) | 1.12 ( 1.05 , 1.20 ) | 0.999 | 37.0 ( 89.7 , 23.2 ) |
|  | North West | 0.025 ( 0.020 , 0.031 ) | 1.17 ( 1.13 , 1.21 ) | 1.000 | 27.2 ( 35.1 , 22.2 ) |
|  | Yorkshire and The Humber | 0.034 ( 0.025 , 0.043 ) | 1.23 ( 1.17 , 1.29 ) | 1.000 | 20.5 ( 27.8 , 16.2 ) |
|  | East Midlands | 0.030 ( 0.023 , 0.038 ) | 1.20 ( 1.15 , 1.26 ) | 1.000 | 22.7 ( 30.5 , 18.1 ) |
|  | West Midlands | 0.041 ( 0.031 , 0.050 ) | 1.28 ( 1.21 , 1.35 ) | 1.000 | 17.0 ( 22.1 , 13.9 ) |
|  | East of England | 0.026 ( 0.017 , 0.035 ) | 1.17 ( 1.11 , 1.24 ) | 1.000 | 26.7 ( 41.1 , 19.7 ) |
|  | London | 0.023 ( 0.014 , 0.033 ) | 1.16 ( 1.09 , 1.22 ) | 1.000 | 29.6 ( 48.7 , 21.1 ) |
|  | South West | 0.029 ( 0.018 , 0.040 ) | 1.19 ( 1.12 , 1.28 ) | 1.000 | 24.1 ( 38.6 , 17.2 ) |
| Round 6a | South East | 0.162 ( 0.052 , 0.271 ) | 2.34 ( 1.36 , 3.61 ) | 0.999 | 4.3 ( 13.4 , 2.6 ) |
|  | North East | -0.079 ( -0.248 , 0.085 ) | 0.57 ( 0.07 , 1.63 ) | 0.170 | -8.8 ( -2.8 , 8.1 ) |
|  | North West | 0.031 ( -0.049 , 0.110 ) | 1.21 ( 0.72 , 1.84 ) | 0.774 | 22.6 ( -14.3 , 6.3 ) |
|  | Yorkshire and The Humber | 0.075 ( -0.026 , 0.174 ) | 1.54 ( 0.84 , 2.47 ) | 0.927 | 9.2 ( -27.0 , 4.0 ) |
|  | East Midlands | 0.057 ( -0.033 , 0.151 ) | 1.40 ( 0.80 , 2.23 ) | 0.894 | 12.1 ( -20.9 , 4.6 ) |
|  | West Midlands | 0.107 ( -0.007 , 0.220 ) | 1.81 ( 0.96 , 2.98 ) | 0.968 | 6.5 ( -104.6 , 3.1 ) |
|  | East of England | 0.146 ( 0.023 , 0.278 ) | 2.18 ( 1.15 , 3.70 ) | 0.990 | 4.7 ( 30.0 , 2.5 ) |
|  | London | 0.210 ( 0.066 , 0.359 ) | 2.86 ( 1.47 , 4.87 ) | 0.998 | 3.3 ( 10.5 , 1.9 ) |
|  | South West | 0.133 ( -0.018 , 0.284 ) | 2.06 ( 0.89 , 3.79 ) | 0.957 | 5.2 ( -38.3 , 2.4 ) |
| Round 6b | South East | -0.018 ( -0.074 , 0.037 ) | 0.89 ( 0.59 , 1.25 ) | 0.264 | -38.3 ( -9.4 , 18.6 ) |
|  | North East | 0.023 ( -0.076 , 0.118 ) | 1.15 ( 0.58 , 1.91 ) | 0.682 | 30.0 ( -9.1 , 5.9 ) |
|  | North West | -0.028 ( -0.083 , 0.022 ) | 0.83 ( 0.55 , 1.15 ) | 0.140 | -24.8 ( -8.4 , 30.9 ) |
|  | Yorkshire and The Humber | -0.019 ( -0.095 , 0.063 ) | 0.88 ( 0.50 , 1.45 ) | 0.322 | -36.6 ( -7.3 , 11.0 ) |
|  | East Midlands | -0.038 ( -0.106 , 0.028 ) | 0.77 ( 0.45 , 1.19 ) | 0.128 | -18.2 ( -6.5 , 24.6 ) |
|  | West Midlands | -0.037 ( -0.104 , 0.028 ) | 0.78 ( 0.46 , 1.18 ) | 0.132 | -18.7 ( -6.6 , 25.1 ) |
|  | East of England | 0.018 ( -0.058 , 0.092 ) | 1.12 ( 0.67 , 1.69 ) | 0.681 | 38.5 ( -12.0 , 7.5 ) |
|  | London | -0.038 ( -0.110 , 0.034 ) | 0.77 ( 0.43 , 1.23 ) | 0.158 | -18.3 ( -6.3 , 20.4 ) |
|  | South West | -0.008 ( -0.091 , 0.073 ) | 0.95 ( 0.51 , 1.53 ) | 0.423 | -84.5 ( -7.6 , 9.5 ) |

**Table 5a.**
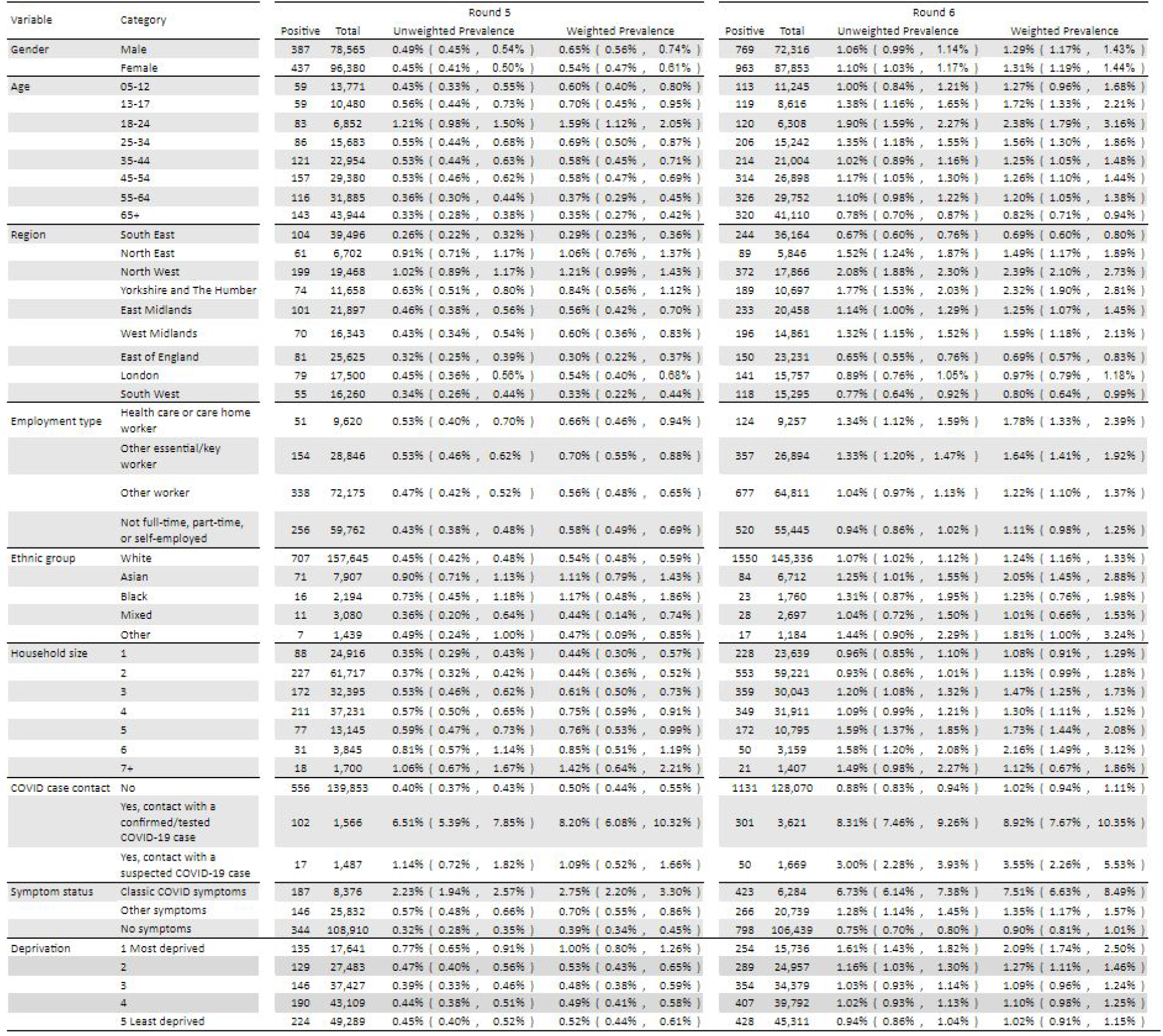
Unweighted and weighted prevalence of swab-positivity by variable and category for rounds 5 and 6.

**Table 5b.** Unweighted and weighted prevalence of swab-positivity by variable and category for rounds 6a and 6b.

| Variable | Category | Round 6a |  |  |  | Round 6b |  |  |  |
| --- | --- | --- | --- | --- | --- | --- | --- | --- | --- |
|  |  | Positive | Total | Unweighted Prevalence | Weighted Prevalence | Positive | Total | Unweighted Prevalence | Weighted Prevalence |
| Gender | Male | 387 | 39,319 | 0.98% ( 0.89% , 1.09% ) | 1.31% ( 1.13% , 1.53% ) | 382 | 33,000 | 1.16% ( 1.05% , 1.28% ) | 1.27% ( 1.11% , 1.45% ) |
|  | Female | 476 | 46,648 | 1.02% ( 0.93% , 1.12% ) | 1.25% ( 1.10% , 1.43% ) | 487 | 41,208 | 1.18% ( 1.08% , 1.29% ) | 1.37% ( 1.20% , 1.57% ) |
| Age | 05-12 | 55 | 5,513 | 1.00% ( 0.77% , 1.30% ) | 1.49% ( 0.98% , 2.26% ) | 58 | 5,732 | 1.01% ( 0.78% , 1.31% ) | 1.07% ( 0.77% , 1.48% ) |
|  | 13-17 | 60 | 5,169 | 1.16% ( 0.90% , 1.49% ) | 1.58% ( 1.08% , 2.31% ) | 59 | 3,447 | 1.71% ( 1.33% , 2.20% ) | 1.92% ( 1.40% , 2.63% ) |
|  | 18-24 | 53 | 3,091 | 1.71% ( 1.31% , 2.24% ) | 2.25% ( 1.47% , 3.42% ) | 67 | 3,217 | 2.08% ( 1.64% , 2.64% ) | 2.51% ( 1.71% , 3.67% ) |
|  | 25-34 | 97 | 7,186 | 1.35% ( 1.11% , 1.64% ) | 1.62% ( 1.25% , 2.08% ) | 109 | 8,056 | 1.35% ( 1.12% , 1.63% ) | 1.50% ( 1.18% , 1.92% ) |
|  | 35-44 | 98 | 10,079 | 0.97% ( 0.80% , 1.18% ) | 1.29% ( 0.98% , 1.68% ) | 116 | 10,925 | 1.06% ( 0.89% , 1.27% ) | 1.22% ( 0.99% , 1.51% ) |
|  | 45-54 | 137 | 13,009 | 1.05% ( 0.89% , 1.24% ) | 1.22% ( 0.99% , 1.51% ) | 177 | 13,889 | 1.27% ( 1.10% , 1.47% ) | 1.29% ( 1.08% , 1.54% ) |
|  | 55-64 | 167 | 15,768 | 1.06% ( 0.91% , 1.23% ) | 1.20% ( 0.99% , 1.46% ) | 159 | 13,984 | 1.14% ( 0.97% , 1.33% ) | 1.21% ( 0.99% , 1.47% ) |
|  | 65+ | 196 | 26,156 | 0.75% ( 0.65% , 0.86% ) | 0.81% ( 0.68% , 0.96% ) | 124 | 14,960 | 0.83% ( 0.70% , 0.99% ) | 0.83% ( 0.67% , 1.03% ) |
| Region | South East | 106 | 19,367 | 0.55% ( 0.45% , 0.66% ) | 0.55% ( 0.45% , 0.68% ) | 138 | 16,797 | 0.82% ( 0.70% , 0.97% ) | 0.84% ( 0.70% , 1.02% ) |
|  | North East | 45 | 3,323 | 1.35% ( 1.01% , 1.81% ) | 1.17% ( 0.84% , 1.64% ) | 44 | 2,524 | 1.74% ( 1.30% , 2.33% ) | 1.88% ( 1.33% , 2.66% ) |
|  | North West | 192 | 10,035 | 1.91% ( 1.66% , 2.20% ) | 2.27% ( 1.90% , 2.72% ) | 180 | 7,831 | 2.30% ( 1.99% , 2.65% ) | 2.53% ( 2.09% , 3.06% ) |
|  | Yorkshire and The Humber | 113 | 6,179 | 1.83% ( 1.52% , 2.19% ) | 2.72% ( 2.12% , 3.50% ) | 76 | 4,518 | 1.68% ( 1.35% , 2.10% ) | 1.80% ( 1.33% , 2.44% ) |
|  | East Midlands | 128 | 11,461 | 1.12% ( 0.94% , 1.33% ) | 1.20% ( 0.98% , 1.47% ) | 105 | 8,997 | 1.17% ( 0.97% , 1.41% ) | 1.31% ( 1.04% , 1.63% ) |
|  | West Midlands | 90 | 8,005 | 1.12% ( 0.92% , 1.38% ) | 1.62% ( 1.05% , 2.49% ) | 106 | 6,856 | 1.55% ( 1.28% , 1.87% ) | 1.56% ( 1.06% , 2.30% ) |
|  | East of England | 74 | 12,612 | 0.59% ( 0.47% , 0.74% ) | 0.64% ( 0.48% , 0.85% ) | 76 | 10,620 | 0.72% ( 0.57% , 0.89% ) | 0.74% ( 0.57% , 0.96% ) |
|  | London | 61 | 7,299 | 0.84% ( 0.65% , 1.07% ) | 0.89% ( 0.66% , 1.19% ) | 80 | 8,459 | 0.95% ( 0.76% , 1.18% ) | 1.03% ( 0.79% , 1.35% ) |
|  | South West | 54 | 7,690 | 0.70% ( 0.54% , 0.92% ) | 0.72% ( 0.52% , 0.99% ) | 64 | 7,608 | 0.84% ( 0.66% , 1.07% ) | 0.88% ( 0.64% , 1.20% ) |
| Employment type | Health care or care home worker | 54 | 4,069 | 1.33% ( 1.02% , 1.73% ) | 2.19% ( 1.35% , 3.54% ) | 70 | 5,212 | 1.34% ( 1.06% , 1.69% ) | 1.47% ( 1.09% , 1.98% ) |
|  | Other essential/key worker | 169 | 12,503 | 1.35% ( 1.16% , 1.57% ) | 1.86% ( 1.46% , 2.36% ) | 188 | 14,427 | 1.30% ( 1.13% , 1.50% ) | 1.46% ( 1.21% , 1.77% ) |
|  | Other worker | 366 | 34,262 | 1.07% ( 0.96% , 1.18% ) | 1.30% ( 1.13% , 1.50% ) | 345 | 33,018 | 1.04% ( 0.94% , 1.16% ) | 1.14% ( 0.97% , 1.35% ) |
|  | Not full-time, part-time, or self-employed | 256 | 33,136 | 0.77% ( 0.68% , 0.87% ) | 0.91% ( 0.76% , 1.08% ) | 228 | 19,547 | 1.17% ( 1.03% , 1.33% ) | 1.41% ( 1.18% , 1.68% ) |
| Ethnic group | White | 793 | 79,406 | 1.00% ( 0.93% , 1.07% ) | 1.24% ( 1.12% , 1.37% ) | 757 | 65,936 | 1.15% ( 1.07% , 1.23% ) | 1.25% ( 1.14% , 1.36% ) |
|  | Asian | 35 | 2,863 | 1.22% ( 0.88% , 1.70% ) | 2.06% ( 1.22% , 3.48% ) | 49 | 3,849 | 1.27% ( 0.96% , 1.68% ) | 2.03% ( 1.29% , 3.19% ) |
|  | Black | 7 | 603 | 1.16% ( 0.56% , 2.38% ) | 1.60% ( 0.72% , 3.52% ) | 16 | 1,157 | 1.38% ( 0.85% , 2.23% ) | 1.02% ( 0.57% , 1.84% ) |
|  | Mixed | 14 | 1,326 | 1.06% ( 0.63% , 1.76% ) | 1.03% ( 0.57% , 1.83% ) | 14 | 1,371 | 1.02% ( 0.61% , 1.71% ) | 0.99% ( 0.53% , 1.82% ) |
|  | Other | 6 | 536 | 1.12% ( 0.51% , 2.42% ) | 1.34% ( 0.54% , 3.31% ) | 11 | 648 | 1.70% ( 0.95% , 3.01% ) | 2.19% ( 1.03% , 4.63% ) |
| Household size | 1 | 106 | 12,368 | 0.86% ( 0.71% , 1.04% ) | 0.92% ( 0.72% , 1.17% ) | 122 | 11,273 | 1.08% ( 0.91% , 1.29% ) | 1.25% ( 0.98% , 1.61% ) |
|  | 2 | 306 | 34,116 | 0.90% ( 0.80% , 1.00% ) | 1.12% ( 0.96% , 1.30% ) | 247 | 25,109 | 0.98% ( 0.87% , 1.11% ) | 1.14% ( 0.91% , 1.41% ) |
|  | 3 | 180 | 15,409 | 1.17% ( 1.01% , 1.35% ) | 1.59% ( 1.24% , 2.04% ) | 179 | 14,634 | 1.22% ( 1.06% , 1.41% ) | 1.35% ( 1.11% , 1.65% ) |
|  | 4 | 161 | 16,329 | 0.99% ( 0.85% , 1.15% ) | 1.36% ( 1.05% , 1.75% ) | 188 | 15,582 | 1.21% ( 1.05% , 1.39% ) | 1.25% ( 1.05% , 1.48% ) |
|  | 5 | 76 | 5,438 | 1.40% ( 1.12% , 1.75% ) | 1.42% ( 1.08% , 1.85% ) | 96 | 5,357 | 1.79% ( 1.47% , 2.18% ) | 2.03% ( 1.58% , 2.61% ) |
|  | 6 | 27 | 1,632 | 1.65% ( 1.14% , 2.40% ) | 2.37% ( 1.40% , 4.00% ) | 23 | 1,527 | 1.51% ( 1.01% , 2.25% ) | 1.94% ( 1.16% , 3.22% ) |
|  | 7+ | 7 | 679 | 1.03% ( 0.50% , 2.11% ) | 0.96% ( 0.39% , 2.34% ) | 14 | 728 | 1.92% ( 1.15% , 3.20% ) | 1.27% ( 0.69% , 2.30% ) |
| COVID case contact | No | 529 | 67,864 | 0.78% ( 0.72% , 0.85% ) | 1.23% ( 1.10% , 1.37% ) | 541 | 54,514 | 0.99% ( 0.91% , 1.08% ) | 1.38% ( 1.24% , 1.54% ) |
|  | Yes, contact with a confirmed/tested COVID-19 case | 147 | 1,787 | 8.23% ( 7.04% , 9.59% ) | 9.56% ( 7.71% , 11.80% ) | 139 | 1,666 | 8.34% ( 7.11% , 9.77% ) | 8.40% ( 6.70% , 10.49% ) |
|  | Yes, contact with a suspected COVID-19 case | 22 | 842 | 2.61% ( 1.73% , 3.92% ) | 4.25% ( 2.16% , 8.19% ) | 28 | 754 | 3.71% ( 2.58% , 5.31% ) | 3.08% ( 1.95% , 4.84% ) |
| Symptom status | Classic COVID symptoms | 201 | 3,381 | 5.95% ( 5.20% , 6.79% ) | 6.91% ( 5.78% , 8.25% ) | 196 | 2,646 | 7.41% ( 6.47% , 8.47% ) | 8.00% ( 6.66% , 9.59% ) |
|  | Other symptoms | 119 | 11,430 | 1.04% ( 0.87% , 1.24% ) | 1.12% ( 0.90% , 1.41% ) | 138 | 8,623 | 1.60% ( 1.36% , 1.89% ) | 1.62% ( 1.32% , 2.00% ) |
|  | No symptoms | 380 | 55,746 | 0.68% ( 0.62% , 0.75% ) | 0.90% ( 0.75% , 1.06% ) | 377 | 45,716 | 0.82% ( 0.75% , 0.91% ) | 0.93% ( 0.81% , 1.07% ) |
| Deprivation | 1 Most deprived | 119 | 7,919 | 1.50% ( 1.26% , 1.80% ) | 2.16% ( 1.66% , 2.81% ) | 135 | 7,819 | 1.73% ( 1.46% , 2.04% ) | 2.01% ( 1.58% , 2.56% ) |
|  | 2 | 143 | 12,750 | 1.12% ( 0.95% , 1.32% ) | 1.32% ( 1.07% , 1.61% ) | 146 | 12,209 | 1.20% ( 1.02% , 1.40% ) | 1.23% ( 1.02% , 1.49% ) |
|  | 3 | 176 | 18,292 | 0.96% ( 0.83% , 1.11% ) | 1.11% ( 0.92% , 1.33% ) | 178 | 16,088 | 1.11% ( 0.96% , 1.28% ) | 1.08% ( 0.91% , 1.28% ) |
|  | 4 | 199 | 21,749 | 0.92% ( 0.80% , 1.05% ) | 1.02% ( 0.86% , 1.21% ) | 208 | 18,043 | 1.15% ( 1.01% , 1.32% ) | 1.21% ( 1.02% , 1.43% ) |
|  | 5 Least deprived | 226 | 25,261 | 0.89% ( 0.79% , 1.02% ) | 0.97% ( 0.83% , 1.14% ) | 202 | 20,051 | 1.01% ( 0.88% , 1.16% ) | 1.09% ( 0.93% , 1.28% ) |

**Figure 5.**
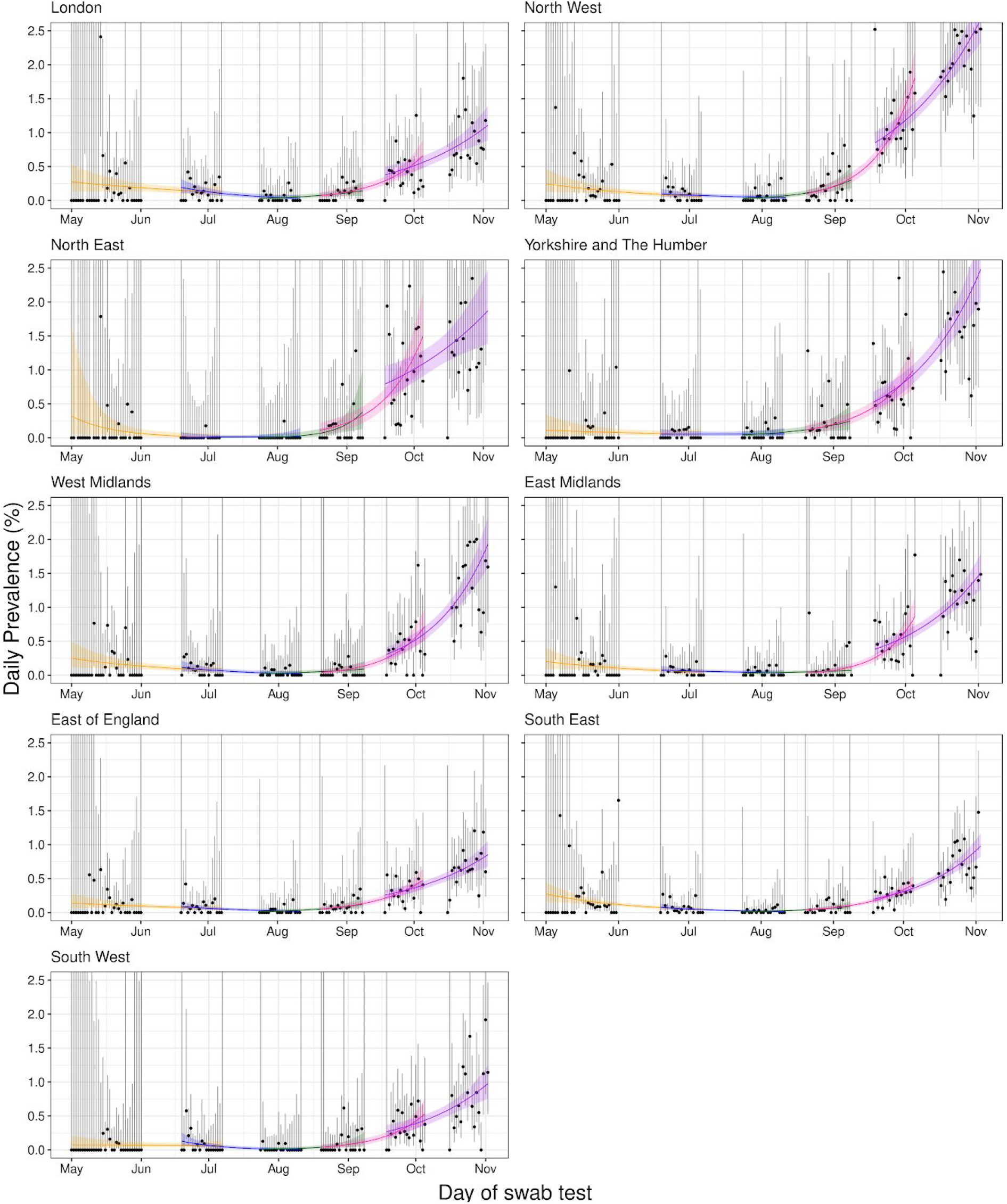
Constant growth rate models fit to regions for REACT-1 data for sequential rounds; 1 and 2 (yellow), 2 and 3 (blue), 3 and 4 (green), 4 and 5 (pink), and 5 and 6 (purple). Points show unweighted prevalence estimate and vertical lines show 95% binomial confidence intervals (CIs). Because there were few swabs taken on some days, some upper bounds to CIs are truncated. Shaded regions show 95% posterior credible intervals for growth models. Note that of the 932,171 swab tests 918,543 had a date and so were included in the temporal analysis (3,008 of 3,029 positives).

**Figure 6.**
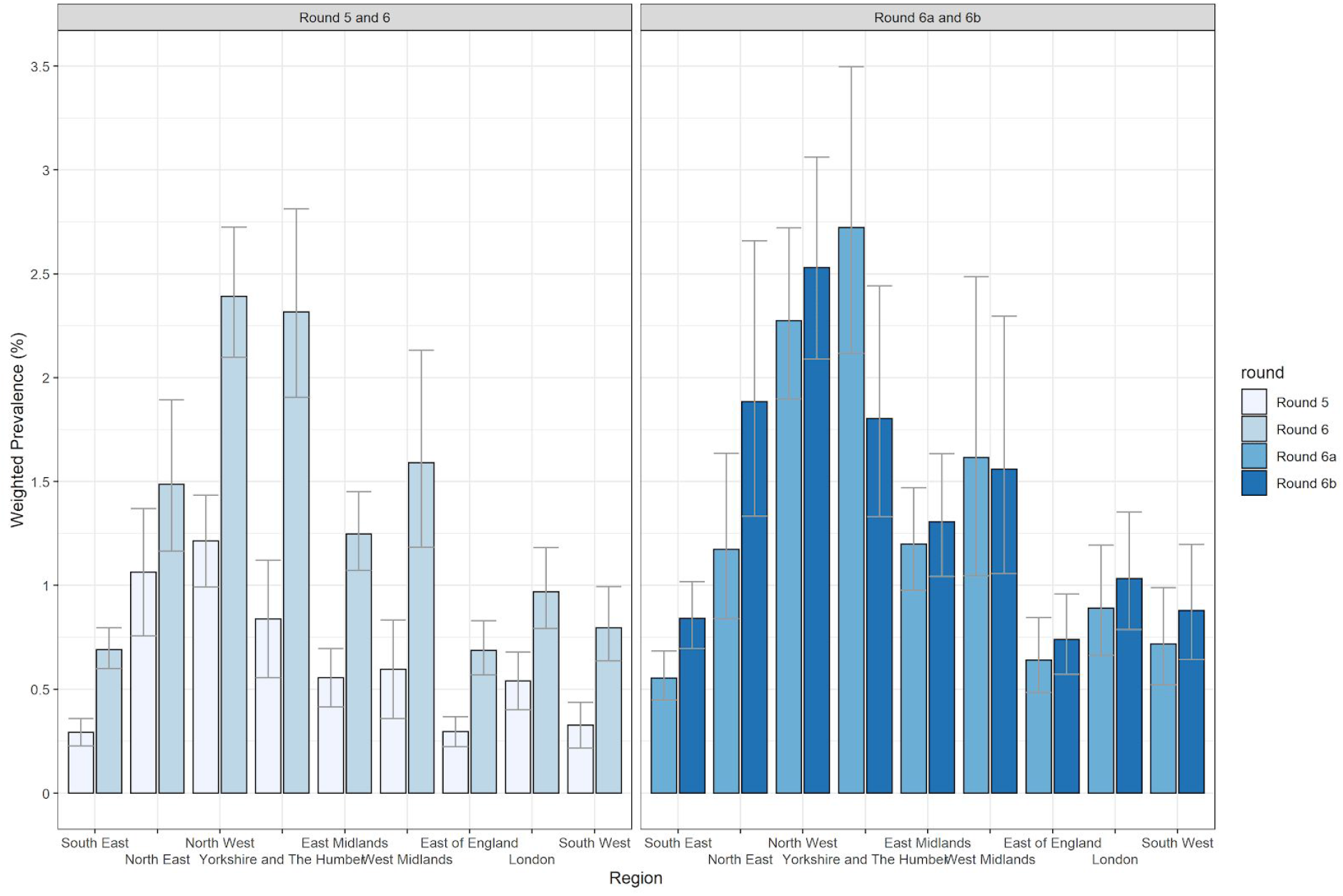
Weighted prevalence of swab positivity by region for rounds 5, 6, 6a and 6b. Bars show 95% confidence intervals.

Differences in local prevalence between rounds 5 and 6, and between the first and second halves of round 6 (averaged at the level of lower tier local authority, LTLA) reveal sub-regional patterns of growth and decline (Figure 7, Figure 8). For example, decline is suggested in a block of LTLAs in the south of Yorkshire and The Humber and the north of East Midlands, while growth is suggested in contiguous blocks of LTLAs across parts of the North West region.

**Figure 7.**
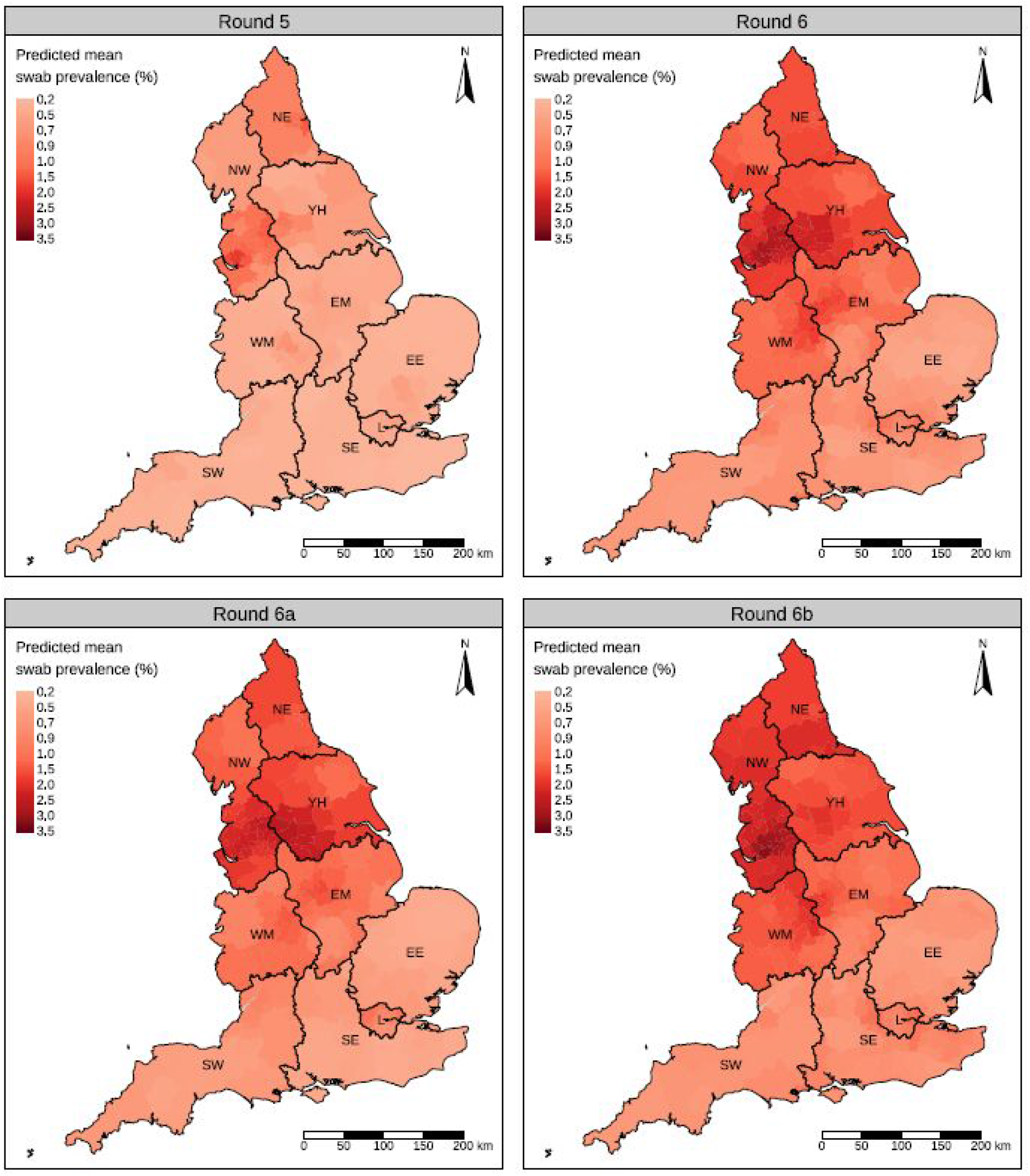
Modelled prevalence at lower tier local authority level (see Methods) for rounds 5, 6, 6a and 6b. Regions: NE = North East, NW = North West, YH = Yorkshire and The Humber, EM = East Midlands, WM = West Midlands, EE = East of England, L = London, SE = South East, SW = South West.

**Figure 8.**
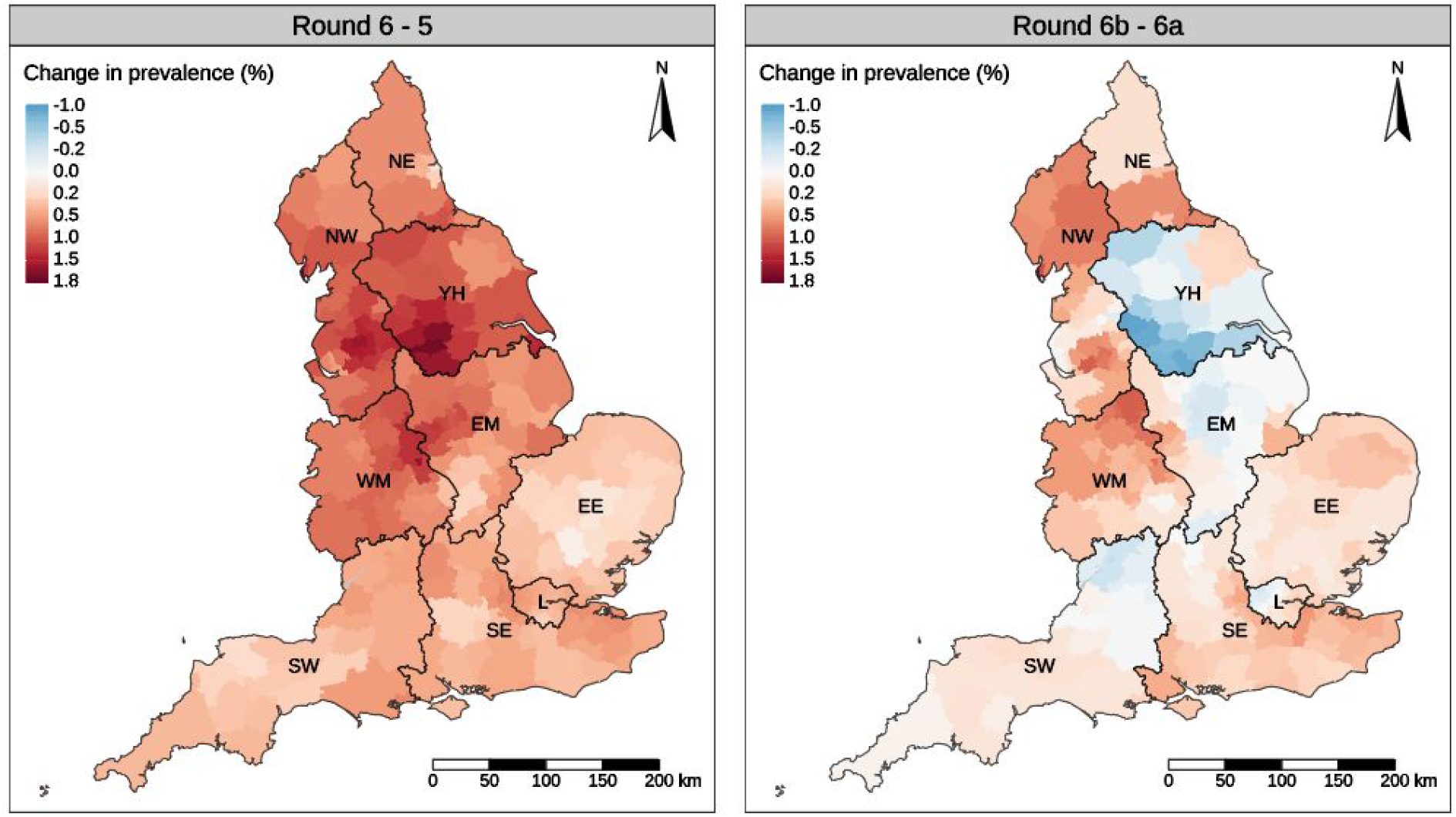
Difference in modelled prevalence at lower tier local authority level (see Methods) between round 6 and round 5, and between rounds 6b and 6a. Regions: NE = North East, NW = North West, YH = Yorkshire and The Humber, EM = East Midlands, WM = West Midlands, EE = East of England, L = London, SE = South East, SW = South West.

In the most recent round 6 data, there is suggestion of decline in weighted prevalence of swab positivity in participants aged 5 to 12 years and to a lesser extent in those aged 25 to 44 years (Table 5b, Figure 9). Again in the most recent data, only age and region were robustly associated with increased odds of testing positive among the covariates, with higher odds of swab positivity in 13-17 and 18-24 year olds and in the North and Midlands compared with the reference groups (Figure 10, Table 6). The estimated odds ratios associated with Black and Asian ethnicity and large household size (six or more individuals per household) were closer to unity in round 6 compared with 5.

**Table 6.** Estimated odds ratios and 95% confidence intervals for jointly adjusted logistic regression model of swab-positivity for rounds 5, 6, 6a (16 to 25 October 2020), and 6b (26 October to 2 November 2020).

| Variable | Category | Round 5 | Round 6 | Round 6a | Round 6b |
| --- | --- | --- | --- | --- | --- |
| Gender | Male | Reference |  |  |  |
|  | Female | 0.89 [0.77,1.03] | 1.01 [0.91,1.11] | 1.03 [0.90,1.19] | 0.98 [0.85,1.13] |
| Age Group | 5 -12 | 0.72 [0.52,1.00] | 0.93 [0.73,1.17] | 1.00 [0.72,1.40] | 0.86 [0.62,1.20] |
|  | 13-17 | 1.01 [0.70,1.47] | 1.54 [1.18,1.99] | 1.63 [1.13,2.35] | 1.59 [1.10,2.30] |
|  | 18-24 | 2.24 [1.68,2.99] | 1.86 [1.48,2.35] | 1.79 [1.27,2.52] | 1.93 [1.41,2.64] |
|  | 25-34 | 1.10 [0.83,1.47] | 1.34 [1.10,1.64] | 1.42 [1.07,1.88] | 1.28 [0.98,1.69] |
|  | 35-44 | Reference |  |  |  |
|  | 45-54 | 1.09 [0.85,1.38] | 1.14 [0.95,1.36] | 1.09 [0.84,1.42] | 1.18 [0.93,1.51] |
|  | 55-64 | 0.79 [0.60,1.03] | 1.14 [0.95,1.37] | 1.21 [0.93,1.58] | 1.09 [0.84,1.42] |
|  | 65+ | 0.76 [0.57,1.02] | 0.93 [0.75,1.14] | 1.04 [0.78,1.39] | 0.86 [0.64,1.16] |
| Region | North East | 3.54 [2.55,4.91] | 2.20 [1.70,2.83] | 2.33 [1.61,3.36] | 2.09 [1.46,2.98] |
|  | North West | 3.91 [3.05,5.02] | 3.11 [2.62,3.68] | 3.51 [2.74,4.49] | 2.80 [2.21,3.54] |
|  | Yorkshire and The Humber | 2.53 [1.86,3.44] | 2.69 [2.21,3.27] | 3.43 [2.61,4.51] | 2.09 [1.56,2.80] |
|  | East Midlands | 1.80 [1.36,2.39] | 1.68 [1.39,2.02] | 2.04 [1.56,2.65] | 1.39 [1.06,1.81] |
|  | West Midlands | 1.65 [1.21,2.25] | 1.86 [1.53,2.27] | 2.09 [1.57,2.79] | 1.68 [1.28,2.21] |
|  | East of England | 1.20 [0.89,1.62] | 0.93 [0.75,1.15] | 1.07 [0.79,1.45] | 0.82 [0.61,1.10] |
|  | London | 1.48 [1.09,2.02] | 1.22 [0.98,1.52] | 1.36 [0.98,1.90] | 1.08 [0.80,1.45] |
|  | South East | Reference |  |  |  |
|  | South West | 1.34 [0.96,1.89] | 1.20 [0.96,1.50] | 1.36 [0.97,1.89] | 1.07 [0.79,1.45] |
| Key Worker Status | Health care or care home worker | 1.03 [0.76,1.39] | 1.14 [0.94,1.40] | 1.12 [0.83,1.50] | 1.15 [0.87,1.50] |
|  | Other essential/key worker | 1.03 [0.85,1.25] | 1.16 [1.01,1.32] | 1.17 [0.97,1.41] | 1.12 [0.94,1.35] |
|  | Other worker | Reference |  |  |  |
|  | Not full-time, part-time, or self-employed | 1.02 [0.84,1.23] | 0.92 [0.81,1.05] | 0.71 [0.60,0.86] | 1.15 [0.95,1.39] |
| Ethnicity | Asian | 1.88 [1.44,2.45] | 1.15 [0.91,1.46] | 1.22 [0.85,1.75] | 1.09 [0.80,1.49] |
|  | Black | 1.67 [1.00,2.78] | 1.31 [0.86,1.99] | 1.24 [0.58,2.64] | 1.32 [0.79,2.20] |
|  | Mixed | 0.77 [0.41,1.44] | 0.94 [0.64,1.40] | 1.02 [0.59,1.78] | 0.89 [0.51,1.55] |
|  | Other | 1.14 [0.54,2.42] | 1.52 [0.94,2.48] | 1.26 [0.56,2.85] | 1.71 [0.93,3.14] |
|  | White | Reference |  |  |  |
| Household Size | 1-2 People | Reference |  |  |  |
|  | 3-5 People | 1.28 [1.08,1.53] | 1.20 [1.07,1.35] | 1.17 [0.99,1.39] | 1.23 [1.04,1.46] |
|  | 6+ People | 1.91 [1.36,2.68] | 1.41 [1.07,1.85] | 1.46 [0.99,2.16] | 1.33 [0.90,1.97] |
| Deprivation Index Quintile | 1 - Most Deprived | Reference |  |  |  |
|  | 2 | 0.80 [0.62,1.03] | 0.87 [0.73,1.04] | 0.88 [0.68,1.13] | 0.87 [0.68,1.12] |
|  | 3 | 0.77 [0.60,0.98] | 0.83 [0.70,0.99] | 0.81 [0.64,1.03] | 0.86 [0.68,1.10] |
|  | 4 | 0.84 [0.66,1.06] | 0.82 [0.69,0.97] | 0.77 [0.61,0.97] | 0.88 [0.69,1.11] |
|  | 5 - Least Deprived | 0.93 [0.74,1.17] | 0.81 [0.69,0.96] | 0.82 [0.65,1.04] | 0.79 [0.62,1.00] |

**Figure 9.**
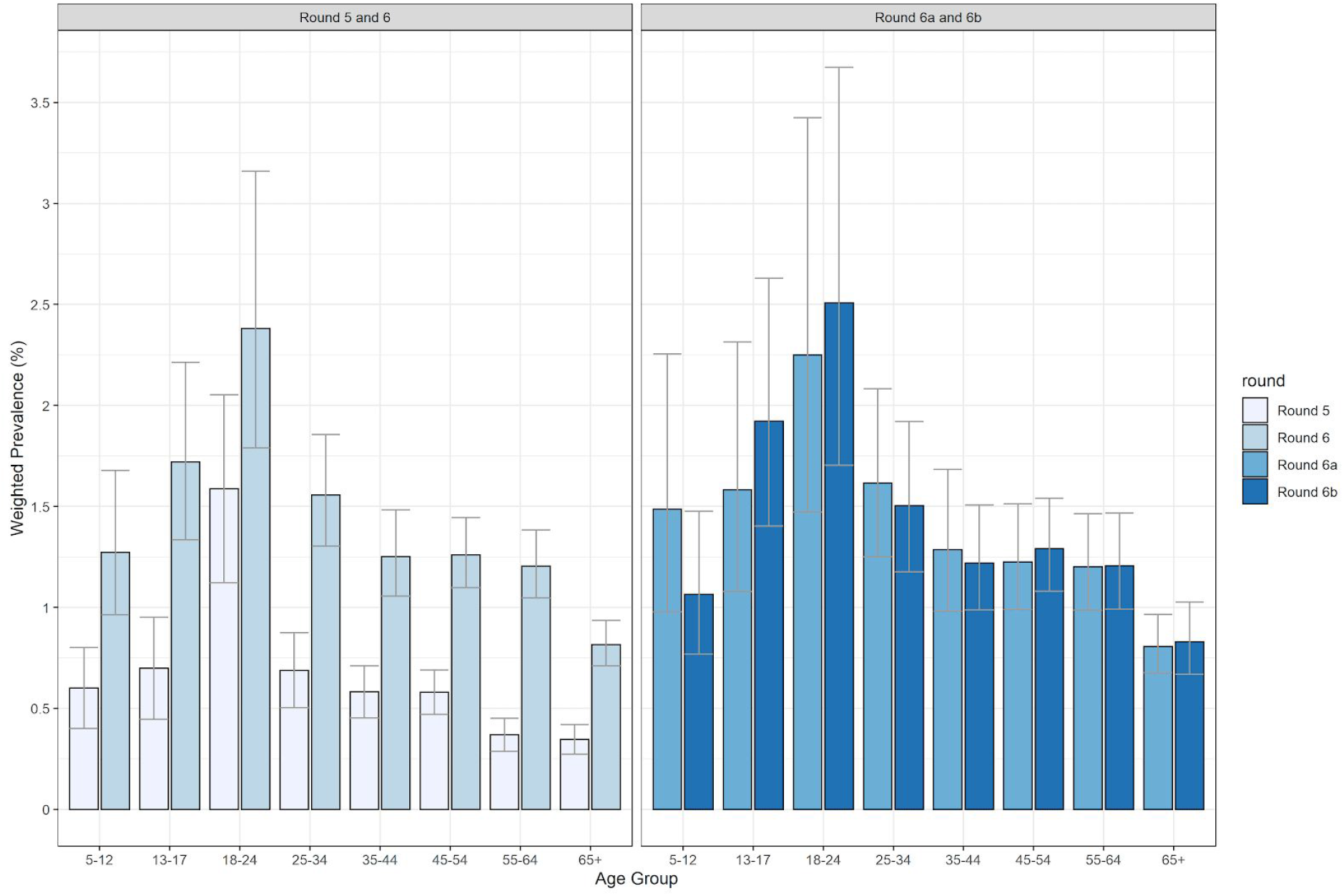
Weighted prevalence and 95% confidence intervals of swab-positivity by age for rounds 5, 6, 6a and 6b.

**Figure 10.**
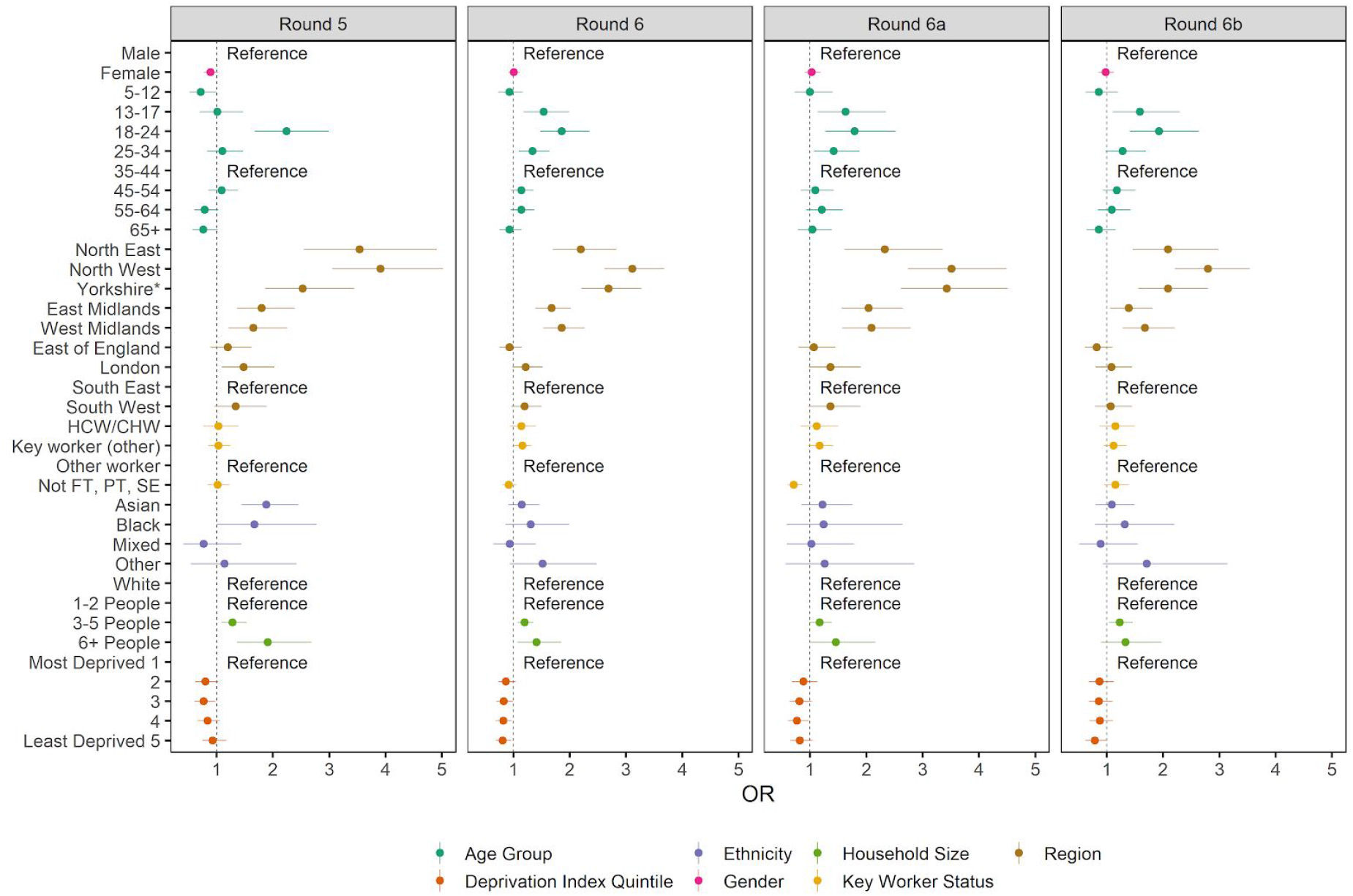
Estimated odds ratios and 95% confidence intervals for jointly adjusted logistic regression model of swab-positivity for rounds 5, 6, 6a (16 to 25 October 2020), and 6b (26 October to 2 November 2020). Models were jointly adjusted for gender, age group, region, key worker status, ethnicity, household size, and deprivation index. The deprivation index is based on the Index of Multiple Deprivation (2019) at lower super output area. Here we group scores into quintiles, where 1 = most deprived and 5 = least deprived. HCW/CHW = health care or care home workers; Not FT, PT, SE = Not full-time, part-time, or self-employed. *Yorkshire and The Humber

## Discussion

During this sixth round of data collection in the REACT-1 study of SARS-CoV-2 virus prevalence in England, we reported high and rapidly increasing prevalence during the first half of the round (16th to 25th October 2020), with highest prevalence in the North of the country [7]. Here we extend these findings to swabs obtained up to 2nd November, three days before a second national lockdown in England. The data presented here therefore give an assessment of community prevalence prior to the second lockdown in England against which to assess the progression of the epidemic during the lockdown period starting 5th November.

In contrast with our findings for mid- to late-October, we found evidence for a slowdown in the epidemic during the final days of October and beginning of November 2020, with suggestion of a fall and then rise in prevalence during that period. This slowdown was seen across the country, both North and South, and was not being driven by any one region. The largest falls in prevalence were seen during late October to beginning November in Yorkshire and The Humber, which had previously had the highest prevalence in the country. We also saw reductions in prevalence at the sub-regional level in that region. Falls in prevalence during this period were also observed at the youngest ages in our study (5 to 12 years).

During this period there was evidence of a downturn in daily infections from the national surveillance data on symptomatic cases (“Pillar 1 and 2”) [1] and from the coronavirus symptom app (Zoe app) [12], and a plateau in data from the Office for National Statistics Coronavirus (COVID-19) Infection Survey [13]. Despite differences between these data streams in their recruitment strategy including whether this is influenced by symptom status [1, 12], all four are broadly consistent in identifying an inflection point towards the end of our study period.

The period leading up to the second national lockdown was fluid, with a number of local initiatives being brought in to curb the virus in the highest prevalence areas, mainly in the North of England and the Midlands. At the same time, there was considerable uncertainty and speculation about a possible national lockdown or whether a “circuit-breaker” might be brought in during half-term week which was also at the end of October for most English local authorities. This reflected concerns that the NHS might not cope with increasing hospital admissions that were already being seen in parts of the country. In addition, October 2020 was one of the wettest Octobers on record in England during which the number of sunshine hours was well below average [14] which may have contributed to changes in behaviour and hence transmission. Overall though, it is difficult to ascribe the patterns of prevalence in the latter part of round 6 to any single cause.

Our national prevalence estimate of 1.3% translates to around 1 million infections in England on any one day, assuming sensitivity to detect the virus from a nose and throat swab of around 75% [15]. If we assume that shedding of the virus is detectable for 10 days on average, this would translate to around 100,000 new infections per day at the end of October with a range from 90,000 to 104,000 (reflecting the 95% confidence intervals in weighted national prevalence).

Our study has a number of limitations. In order to estimate trends over time, we assume that the individuals taking part are broadly representative of the base population by LTLA at each time point in the study. We did find evidence for limited differences in population characteristics between the first and second halves of rounds four to six, but not of sufficient size or direction to have materially affected within- or between-round trends. Also it is possible that issues with swab sample transport or changes in laboratory procedures or reagents may have affected either the integrity of the samples or the detection thresholds on RT-PCR. However, data are collected and analysed according to strict protocols and quality control (QC) procedures, and review of both the delivery chain and laboratory QC did not reveal any differences or discrepancies that might have materially affected positivity rates.

Underlying the national trends, we have described a complex spatial pattern of growth and decline of the epidemic at sub-regional scales in the most recent data. Increased restrictions in west Yorkshire prior to the start of the national lockdown [8] appear to have been successful. In the North West, while there were signs of slowing of the epidemic in the worst affected areas, we did not see substantial groupings of LTLAs with lower prevalence in the second half of round 6 compared with the first. These differences suggest variation in the efficacy of the tiered local interventions which may reflect differences in timing, implementation or population acceptability and adherence to the restrictions.

The impact of the second national lockdown in England is not yet known. We provide here a detailed description of swab-positivity patterns at national, regional and local scales for the period immediately preceding lockdown, against which future trends in prevalence can be evaluated.

## Data Availability

The datasets generated or analysed, or both, during this study are not publicly available because of governance restrictions.

## Acknowledgements

SR, CAD acknowledge support: MRC Centre for Global Infectious Disease Analysis, National Institute for Health Research (NIHR) Health Protection Research Unit (HPRU), Wellcome Trust (200861/Z/16/Z, 200187/Z/15/Z), and Centres for Disease Control and Prevention (US, U01CK0005-01-02). GC is supported by an NIHR Professorship. PE is Director of the MRC Centre for Environment and Health (MR/L01341X/1, MR/S019669/1). PE acknowledges support from Health Data Research UK (HDR UK), the NIHR Imperial Biomedical Research Centre and the NIHR HPRUs in Environmental Exposures and Health and Chemical and Radiation Threats and Hazards, the British Heart Foundation Centre for Research Excellence at Imperial College London (RE/18/4/34215) and the UK Dementia Research Institute at Imperial (MC_PC_17114). We thank The Huo Family Foundation for their support of our work on COVID-19.

We thank key collaborators on this work -- Ipsos MORI: Kelly Beaver, Sam Clemens, Gary Welch, Nicholas Gilby, Andrew Cleary and Kelly Ward; Institute of Global Health Innovation at Imperial College: Gianluca Fontana, Dr Hutan Ashrafian, Sutha Satkunarajah and Lenny Naar; MRC Centre for Environment and Health, Imperial College London: Daniela Fecht; North West London Pathology and Public Health England for help in calibration of the laboratory analyses; NHS Digital for access to the NHS register; and the Department of Health and Social Care for logistic support. SR acknowledges helpful discussion with attendees of meetings of the UK Government Office for Science (GO-Science) Scientific Pandemic Influenza – Modelling (SPI-M) committee.

## Tables and Figures

Supporting data to support tables and figures are available [here](https://docs.google.com/spreadsheets/d/10C9gX6RSnEoXf0h9HkbtqEyo7WqR4MQx7nAhhvNiL4k/edit?usp=sharing).

